# Large-scale validation of the Prediction model Risk Of Bias ASsessment Tool (PROBAST) using a short form: high risk of bias models show poorer discrimination

**DOI:** 10.1101/2021.01.20.21250183

**Authors:** Esmee Venema, Benjamin S Wessler, Jessica K Paulus, Rehab Salah, Gowri Raman, Lester Y Leung, Benjamin C Koethe, Jason Nelson, Jinny G Park, David van Klaveren, Ewout W Steyerberg, David M Kent

## Abstract

**Objective:** To assess whether the Prediction model Risk Of Bias ASsessment Tool (PROBAST) and a shorter version of this tool can identify clinical prediction models (CPMs) that perform poorly at external validation.

**Study Design and Setting:** We evaluated risk of bias (ROB) on 102 CPMs from the Tufts CPM Registry, comparing PROBAST to a short form consisting of six PROBAST items anticipated to best identify high ROB. We then applied the short form to all CPMs in the Registry with at least 1 validation and assessed the change in discrimination (dAUC) between the derivation and the validation cohorts (n=1,147).

**Results:** PROBAST classified 98/102 CPMS as high ROB. The short form identified 96 of these 98 as high ROB (98% sensitivity), with perfect specificity. In the full CPM registry, 529/556 CPMs (95%) were classified as high ROB, 20 (4%) low ROB, and 7 (1%) unclear ROB. Median change in discrimination was significantly smaller in low ROB models (dAUC −0.9%, IQR −6.2%–4.2%) compared to high ROB models (dAUC −11.7%, IQR −33.3%–2.6%; p<0.001).

**Conclusion:** High ROB is pervasive among published CPMs. It is associated with poor performance at validation, supporting the application of PROBAST or a shorter version in CPM reviews.

**What is new:**

- High risk of bias is pervasive among published clinical prediction models
- High risk of bias identified with PROBAST is associated with poorer model performance at validation
- A subset of questions can distinguish between models with high and low risk of bias

## 1.0 Introduction

A clinical prediction model (CPM) estimates an individual’s probability of having a disease =(diagnosis) or develop a clinical outcome (prognosis) based on the combination of relevant characteristics. Such models enable researchers or clinicians to inform individuals about their expected outcome and can be used to select patients for a certain treatment or study.^1-3^ The development of a CPM consists of several important steps, including careful predictor selection and model specification.^4, 5^ Methodological shortcomings may cause bias and systematic overestimation of the model performance measures, promoting misleading conclusions of its value for clinical practice.

The Prediction model Risk Of Bias ASsessment Tool (PROBAST) was developed to assess risk of bias (ROB) based on the methods used for model development.^8^ It was developed for systematic reviews and provides a comprehensive overview of methodological quality. It is unclear whether adherence to the methodological standards of PROBAST is associated with a better performance in external validation. Moreover, PROBAST requires both subject and methodological expertise to apply and might be too time intensive for large-scale use. We aimed to examine whether poor PROBAST scores can identify CPMs that perform poorly at external validation and to develop a short form that is equally capable to identify poorly performing CPMs.

## 2.0 Methods

We used publications from the Tufts Predictive Analytics and Comparative Effectiveness (PACE) CPM Registry, a database which includes CPMs for cardiovascular disease published in English language from January 1990 through March 2015 (www.pacecpmregistry.org). A systematic literature search was performed to comprehensively identify CPMs.^9, 10^ For this registry, a de novo CPM was defined as a newly derived prediction model to estimate an individual patient’s absolute risk for a binary outcome. Information from each CPM was extracted from the original article and entered in the database. CPMs were characterized based on the index condition, including coronary artery disease, congestive heart failure, arrhythmias, stroke, venous thromboembolism, and peripheral vascular disease. Populations at risk for developing incident CVD were classified with the index condition of ‘population sample’.

### 2.1 External validations

We included validations of each de novo CPM in the CPM Registry, which have been previously identified with a Scopus citation search conducted on March 22, 2017.^11^ An external validation was defined as any model evaluation on a dataset distinct from the derivation data, including validations that were performed on a temporally or geographically distinct part of the same cohort (i.e., non-random split sample), that reported at least one measure of model performance (discrimination and/or calibration). Discrimination, expressed as the area under the receiver operating characteristic curve (AUC), describes how well a model separates those who develop the outcome of interest from those who do not. Calibration refers to the agreement between the observed and predicted probabilities.

To assess clinical relatedness of the derivation and validation populations, relatedness rubrics were constructed for the 10 most common index conditions and included details on the population sample, major inclusion and exclusion criteria, outcome measure, enrolment period, type of intervention, and follow-up duration. Details of these rubrics are available elsewhere.^11^ Items were extracted from the CPM Registry and, if necessary, the original articles. The relatedness rubrics were used to distinguish between ‘related’ populations, with a near exact match on relevant items for each specific index condition, and ‘distantly related’ populations. Validation cohorts with a different index condition were excluded from analysis.

### 2.2 PROBAST assessment

The PROBAST tool assesses risk of bias based on 20 signalling questions in 4 key domains: participants (e.g. study design and patient inclusion), predictors (e.g., differences in predictor definitions), outcome (e.g., differences in outcome assessment), and analyses (e.g., sample size and handling of missing data). All questions are phrased so that “yes” indicates low ROB and “no” high ROB. As a result, a domain where all questions are answered with “yes” is rated low ROB, while questions answered with “no” indicate a high ROB. Unclear ROB is assigned to a domain when there is insufficient information to assess one or more questions. The overall judgment is considered low ROB when all domains have low ROB. If at least 1 domain has high ROB, the overall judgment is high ROB as well. If at least 1 domain has unclear ROB and all other domains have low ROB, the overall judgment is unclear ROB.^8, 12^

We applied the PROBAST tool to all stroke models with at least one external validation and a reported derivation AUC (n=52) and 50 randomly selected models with other index conditions. Each of these 102 models was assessed by two independent reviewers—blinded to validation study results--using the guidelines in the PROBAST ‘Explanation and Elaboration’.^12^ Discrepancies were discussed with a third reviewer to arrive at a consensus.

### 2.3 Short form

For practical reasons, a selection was made from items in the original PROBAST. We discussed the relevance of all 20 questions within a group of methodologists with specialized expertise in prediction (including DvK, EWS, and DMK). We rated items according to their potential effect on performance of a CPM at validation. We aimed for a subset of questions that can be applied by a trained research assistant (i.e., trained study staff without doctoral-level clinical or statistical expertise) within 15 minutes. Usability of the items was verified through testing by experienced research assistants at Tufts PACE Center.

The following 6 items were considered most relevant and easy to use: ‘outcome assessment’, ‘events per variable (EPV)’, ‘continuous predictors’, ‘missing data’, ‘univariable analysis’, and ‘correction for overfitting/optimism’. We adjusted the definitions from the original PROBAST article to improve clarity and resolve unambiguity (see Supplemental Material). One point was assigned for each item that was incorrectly performed, resulting in total scores ranging from 0 to 6 points. As with the full PROBAST, models with a total score of 0 were classified as ‘low ROB’ and models with a score ≥1 as ‘high ROB’. When the total score was 0 but there was insufficient information provided to assess all items, the model was rated ‘unclear ROB’. We assumed that the effect of using univariable selection or not correcting for optimism would be negligible when the effective sample size was very large. Hence, we did not assign points to these items when the EPV was ≥25 for candidate predictors, or ≥50 for the final model (when candidate predictors were unknown).

We applied this short form to the same set of 102 models and compared results with those of the full PROBAST. We then refined and clarified the scoring guidelines. Research assistants of Tufts PACE center then applied the short form to all de novo CPMs with at least one external validation in the Registry (n=556). Blinded double assessment of the first 40 models was done to compare the assessments and discuss discrepancies. Because the short form was composed of a subset of PROBAST items, CPMs classified as high ROB by the short form were anticipated to also be classified as high ROB by the full PROBAST. CPMs classified by the short form as low ROB might be reclassified as high ROB by the full PROBAST. Thus, all models that were rated as low or unclear ROB were re-assessed by a separate reviewer using PROBAST to reveal any potential items suggestive of high ROB not captured by the short form.

### 2.4 Analyses

Cohen’s kappa statistic was calculated to assess interrater reliability and agreement between PROBAST and the short form. As a measure of the observed ROB in each derivation-validation pair, we used the change in discriminative performance between the derivation and validation cohorts, as quantified by the AUC. The AUC ranges from 0.5 (similar to a coin flip) to 1.0 (perfect discrimination).^13^ The percent change in discrimination was thus calculated:

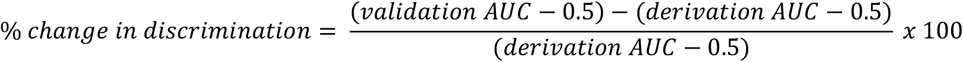

For example, when the AUC decreases from 0.70 in derivation to 0.60 in validation, the delta AUC (dAUC) represents a 50% loss in discriminative ability (since 0.50 is the reference value for AUC). We calculated the median and interquartile range (IQR) of the change in discrimination for low ROB versus high ROB models and stratified for relatedness.

We used generalized estimation equations (GEE) with robust covariance estimator^14, 15^ to assess the association between the ROB classification and the observed change in discrimination, taking into account the correlation between validations of the same CPM. In these analyses, we calculated the absolute difference between two dAUCs. For example, if one model had dAUC 20%, while the other model had a dAUC 6%, the difference in dAUC would be 14%. We also constructed a multivariable GEE model to control for the following factors: relatedness, index condition, CPM authors (same as in validation paper, author overlap, no author overlap), CPM method (logistic, time-to-event, other), CPM center (single, multicenter), CPM source (medical record, registry, trial, other), validation design (cohort, trial, other), validation center (single, multicenter), validation source (medical record, registry, trial, other), CPM parameter degrees of freedom, CPM events per variable, CPM sample size, CPM events, validation events per variable, validation sample size, validation events, relative outcome rate difference >40%, and difference in years between CPM and validation. For this multivariable model, multiple imputation using 20 imputation sets was used to account for missing data. All statistical analyses were performed using SAS Enterprise Guide version 8.2 (SAS Institute Inc., Cary, NC, USA).

## 3.0 Results

PROBAST was assessed on the first set of 102 models (52 stroke models and 50 models). Of these models, 98 (96%) were rated high ROB and only 4 (3.9%) low ROB. Overall high ROB was mainly caused by high ROB in the analysis domain, while the other three domains contributed little information (Figure 1A). Agreement between the two reviewers before the final consensus meeting was 90% for the overall judgment (kappa 0.33). Interrater agreement ranged between 49 and 97% per item (kappa −0.05 to 0.88., Supplemental Table S1). When applying the short form to the same 102 models, the sensitivity to detect high ROB was 98% (using the full PROBAST as reference standard) and specificity was 100%. Overall agreement was good (98%, kappa 0.79). The item ‘outcome assessment’ was rated high ROB in only 4% of the models, while the percentage high ROB of the other items ranged between 39 and 77% (Figure 1B). Figure 2 shows the distribution of the short form total scores.

**Figure 1.**
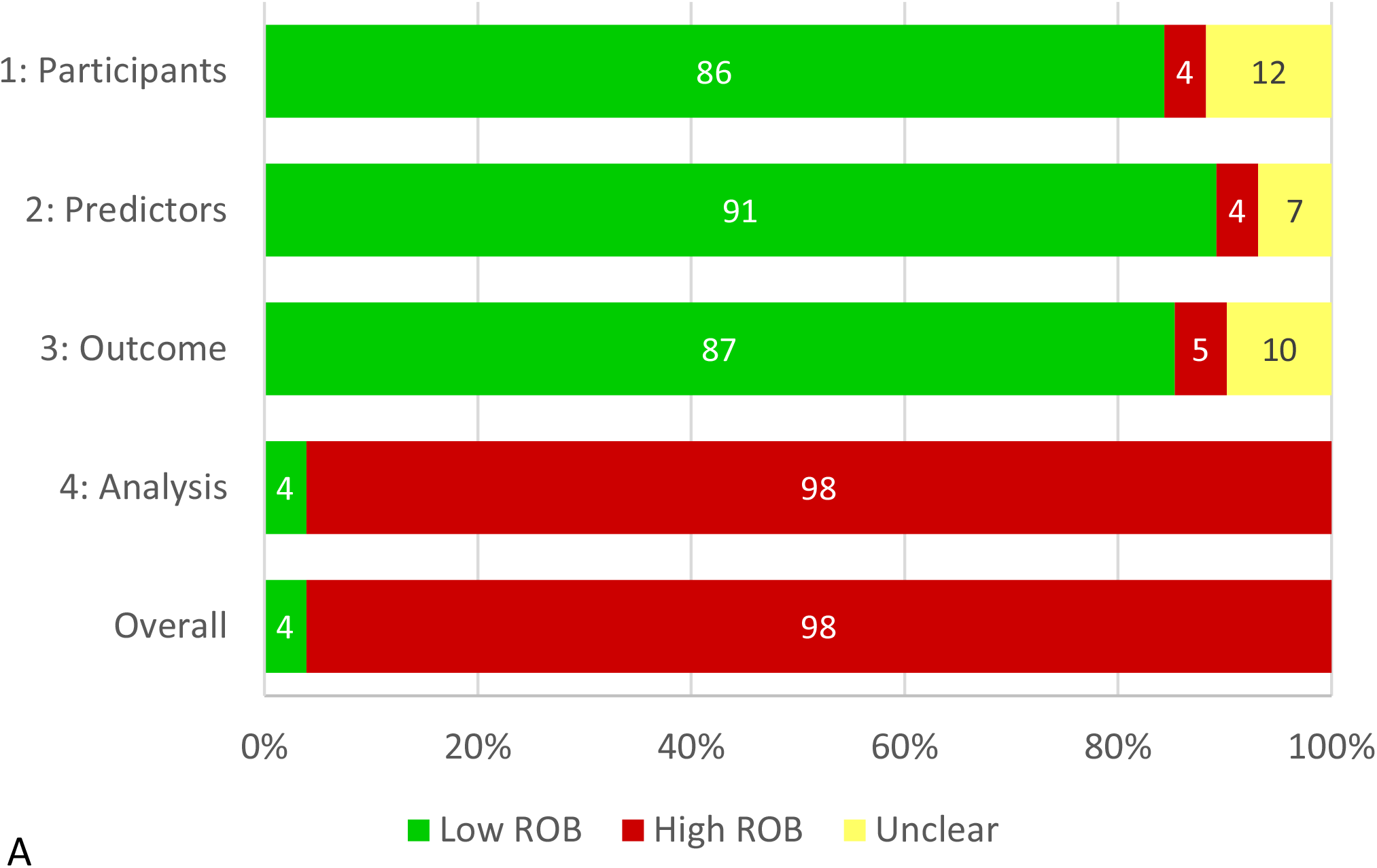
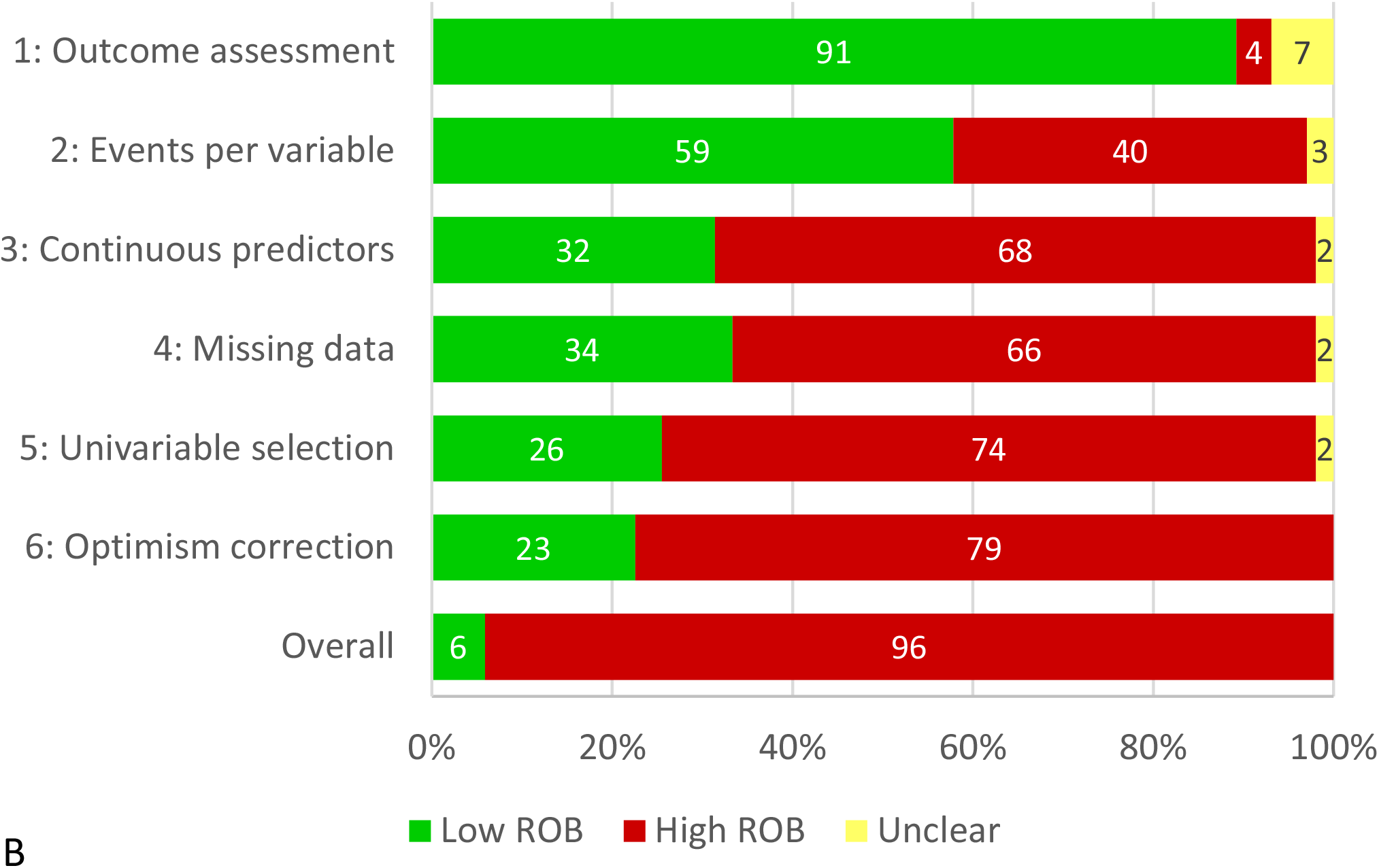
Risk of bias assessed with PROBAST (A, per domain) and the short form (B, per item) in the initial set of 102 clinical prediction models. The first item of the short form (“Outcome assessment”) belongs to domain 3 (“Outcome”), all other items belong to domain 4 (“Analysis”).

**Figure 2.**
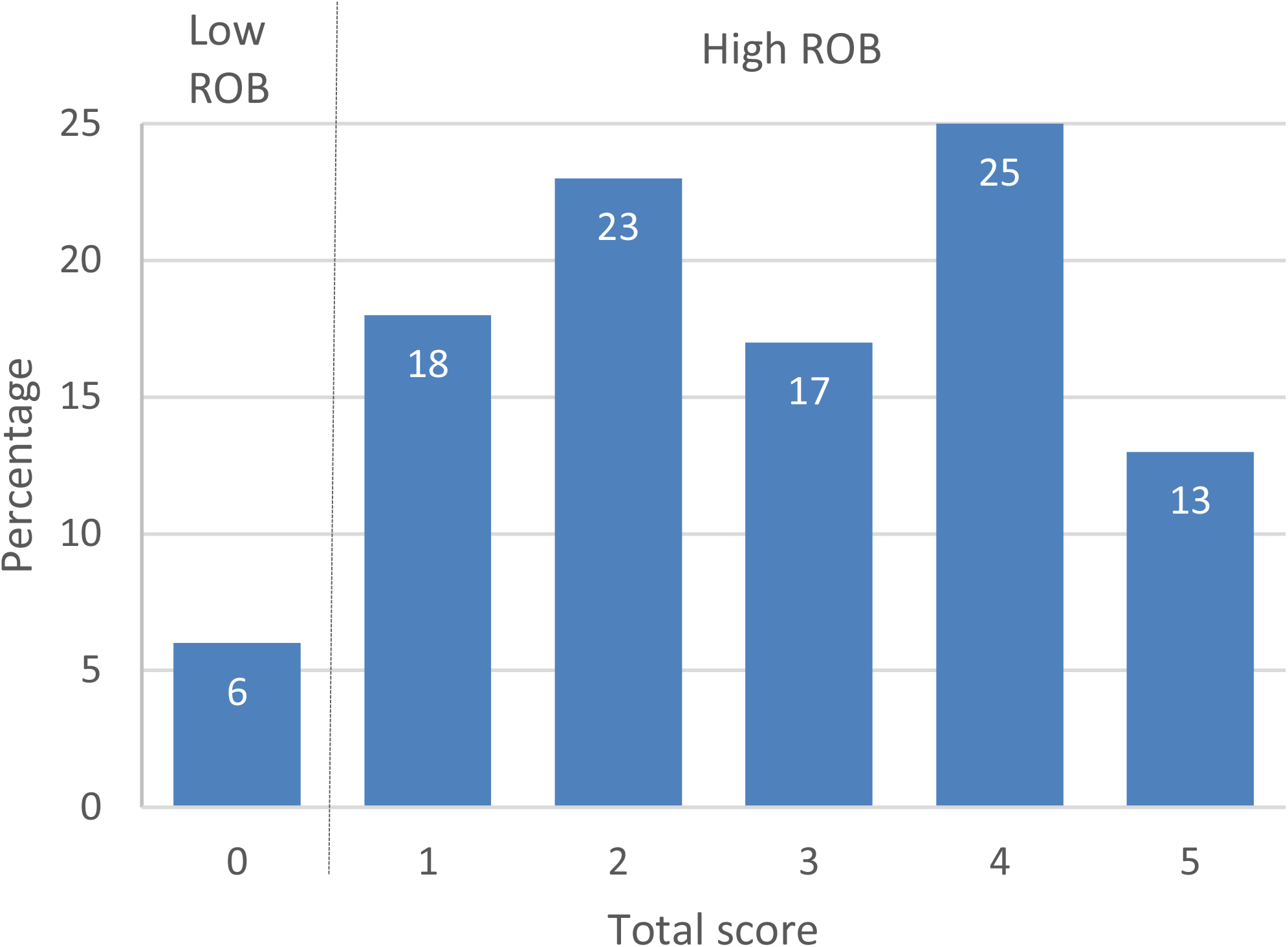
Distribution of the short form total scores in the initial set of 102 clinical prediction models. Total score can range from 0 to 6, with 0 points indicating low risk of bias and ≥1 high risk of bias.

The CPM Registry included 1,382 de novo CPMs, of which 556 (40%) were externally validated at least once. The most common index conditions were coronary artery disease (n=129), stroke (n=97), population sample (n=85), and cardiac surgery (n=70). In total, 1,846 validations of the 556 CPMs were included in the CPM Registry. The number of validations per model ranged from 1 to 86 with a median of 1 (IQR 1 – 3). Relatedness was assessed for 1,702 validations (i.e., of CPMs for the top 10 index conditions), of which 985 (58%) were related and 717 (42%) were distantly related.

Following further clarifications of guidance (see Supplemental Material), the short form was applied to all 556 de novo models in the dataset. In total, 529 (95%) were considered high ROB, 20 (4%) low ROB, and 7 (1%) unclear ROB. Only one model with unclear ROB was reclassified to high ROB after full PROBAST assessment of all low and unclear ROB models. Information on both the derivation AUC and validation AUC was available for 1,147 validations (62%). The median dAUC of the derivation-validation pairs was −10.7% (IQR −32.4% – 2.8%). The difference was significantly smaller in low ROB models (dAUC −0.9%, IQR −6.2% – 4.2%) compared to high ROB models (dAUC −11.7%, IQR −33.3% – 2.6%; p<0.001; Table 1 and Figure 3).

**Table 1.** Percent change in discrimination between derivation AUC and validation AUC (dAUC).

|  | **N** | **N missing** | **N included** | **Median dAUC (IQR)** | **Range** |
| --- | --- | --- | --- | --- | --- |
| **All validations** | **1846** | **699** | **1147** | **-10.7%**<br>**(-32.4% – 2.8%)** | **-179% to 191%** |
| High ROB | 1780 | 667 | 1113 | -11.7%<br>(-33.3% – 2.6%) | -179% to 191% |
| Low ROB | 39 | 11 | 28 | -0.9%<br>(-6.2% – 4.2%) | -20% to 45% |
| Unclear ROB | 27 | 21 | 6 | -13.3%<br>(-25.7% – -4.1%) | -42% to 0% |
| **Related validations** | **985** | **362** | **623** | **-8.3%**<br>**(-25.2% – 3.8%)** | **-129% to 157%** |
| High ROB | 947 | 348 | 599 | -8.8%<br>(-26.2% – 3.8%) | -129% to 157% |
| Low ROB | 28 | 8 | 20 | -1.4%<br>(-6.2% – 3.6%) | -20% to 11% |
| Unclear ROB | 10 | 6 | 4 | -8.6%<br>(-13.3% – -2.1%) | -14% to 0% |
| **Distantly related validations** | **717** | **286** | **431** | **-17.2%**<br>**(-42.1% – 0.2%)** | **-133% to 191%** |
| High ROB | 692 | 269 | 423 | -17.4%<br>(-42.4% – 0%) | -133% to 191% |
| Low ROB | 8 | 2 | 6 | 4.2%<br>(3.3% – 27.3%) | -12% to 31% |
| Unclear ROB | 17 | 15 | 2 | -33.9%<br>(-42.1% – -25.7%) | -42% to -26% |

Relatedness was assessed using relatedness rubrics specifically developed for this purpose.<sup>11</sup> AUC indicates area under the receiver operator characteristic curve; IQR interquartile range; ROB, risk of bias.

**Figure 3.**
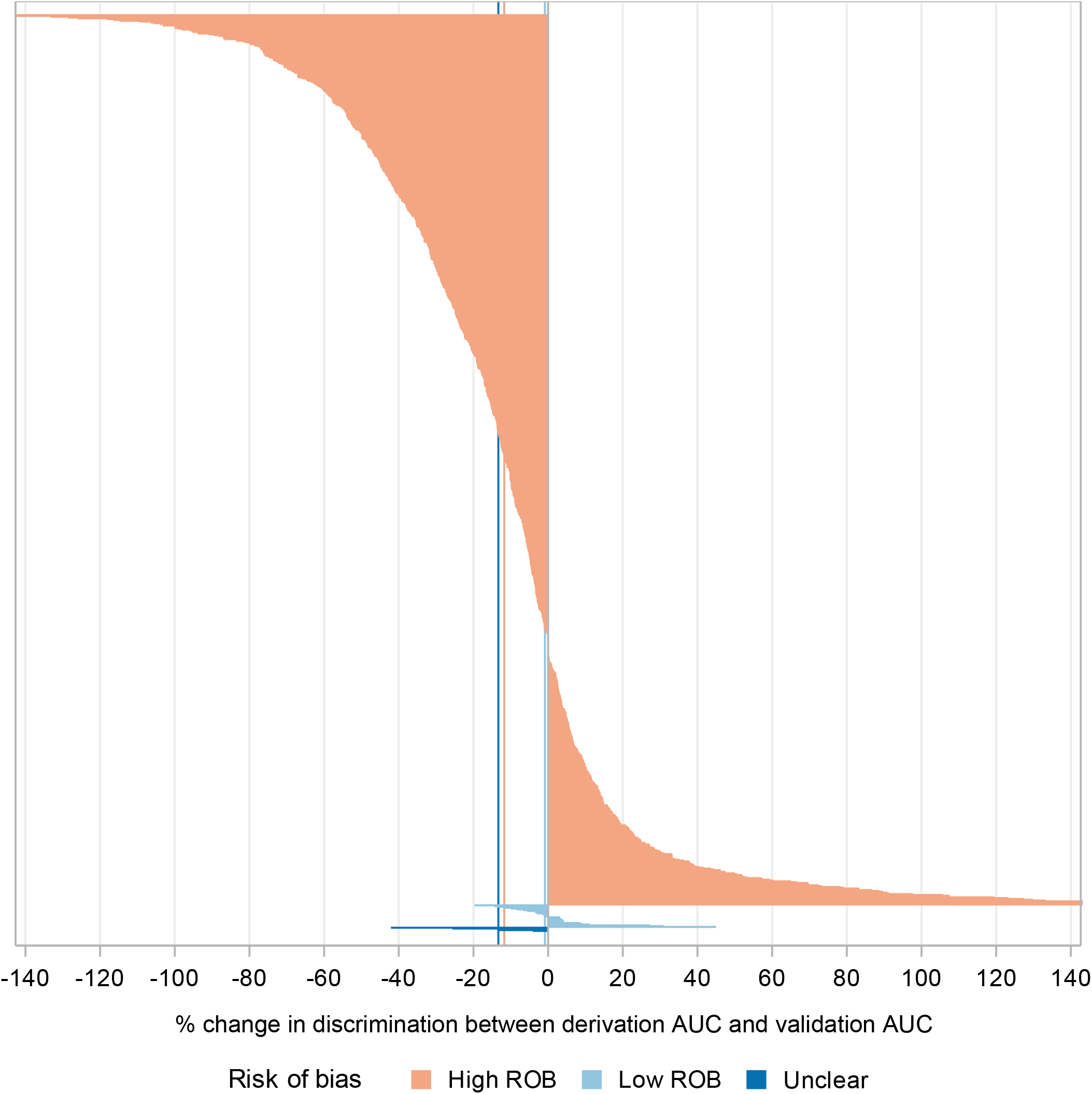
Waterfall plot showing the distribution of delta AUC in each derivation-validation pair (n=1,147), divided by the risk of bias. AUC indicates area under the receiver operating characteristic curve; ROB, risk of bias.

The GEE analyses estimated a difference in dAUC of 16.8% (95% CI 6.1% to 27.6%, p=0.002) for low ROB versus high ROB after adjustment for CPM and validation characteristics (Table 2). This number can be interpreted as an absolute difference between the validation AUC of a high ROB model and a low ROB model of 0.02 when the derivation AUC was 0.60 or as 0.07 when the derivation AUC was 0.90 (Supplemental Table S2).

**Table 2.**
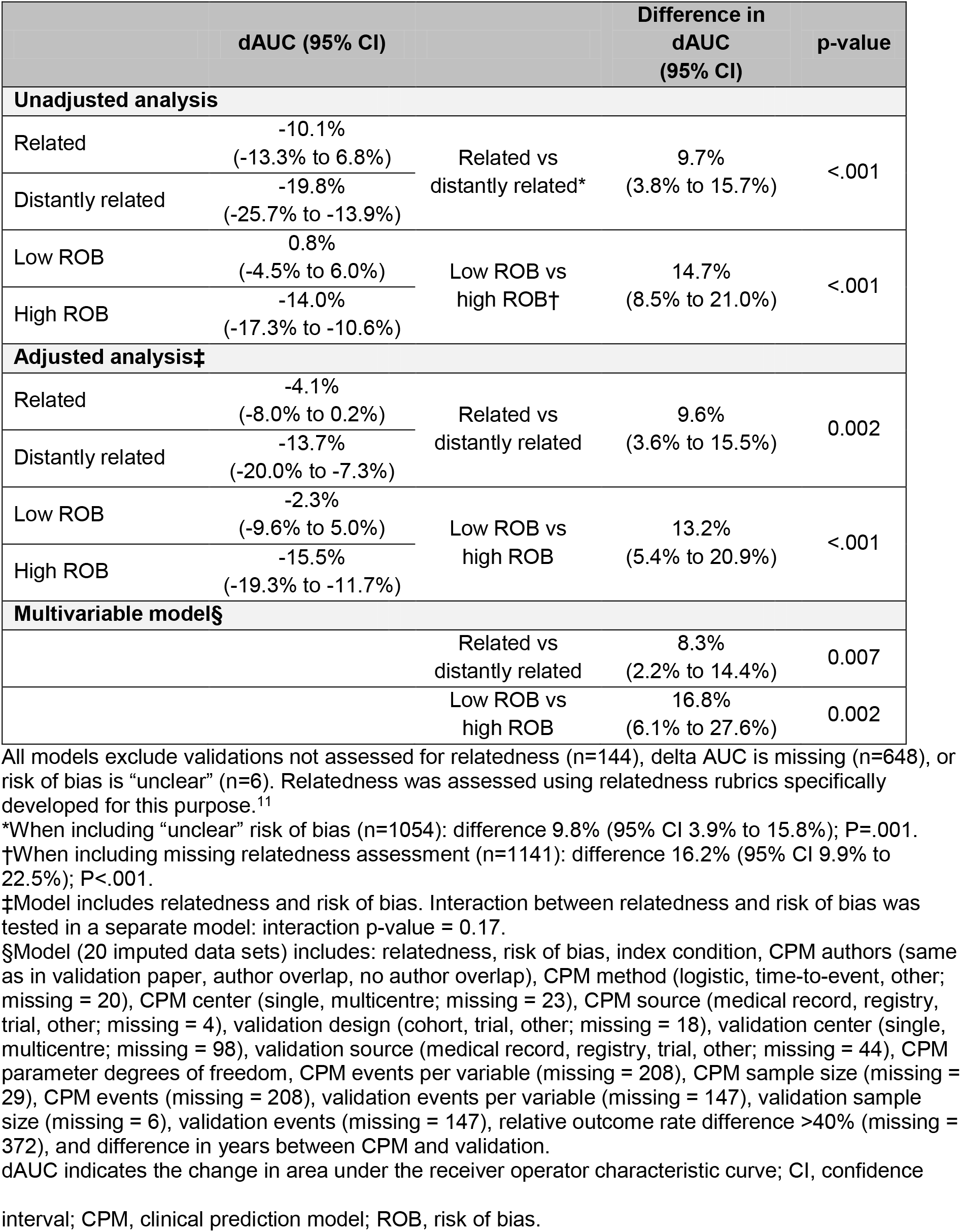
Results of generalized estimated equations (GEE) performed with data of 1048 validations

## 4.0 Discussion

PROBAST can be used to obtain a comprehensive overview of important methodological elements of CPM studies. A selection of six key items from the original 20 items fully captured the overall ROB assessment. We assessed a large set of CPMs of which most models were classified as high ROB, which was associated with poorer discriminative ability at external validation compared to CPMs with low ROB scores.

While we did not formally conduct time assessments, applying the full PROBAST took up to one hour and required both subject and methodological expertise. While some items may be more likely to cause or identify optimism in model performance than others, the tool does not prioritize items. For example, a key pitfall such as using a small dataset with an inadequate number of events-per-variable is weighted the same as not reporting all relevant performance measures. Thus, in constructing a short form, we prioritized those items felt to be most relevant for poor performance at validation. Moreover, while the ‘Explanation and Elaboration’ article provides extensive guidelines for the assessment,^12^ it nevertheless leaves some questions open for interpretation, complicating assessment by research assistants without specific training. We clarified guidelines to diminish ambiguity and reduce interrater variability. The average time of applying the selected subset of 6 questions was approximately 15 minutes.

The selection of items for the short form, which was done strictly by expert opinion and prioritized items in the analysis domain, showed concurrent validity with the PROBAST assessment in 102 papers. Nearly all high ROB CPMs were identified based on violating any item from the analysis domain, which was well represented in the short form. Additionally, because the short form is comprised of a subset of the PROBAST questions, it is 100% specific for identifying low ROB CPMs. Because the number of low ROB CPMs is very small (typically <5% of the total), review of this subset with the full PROBAST should result in identical classification as using PROBAST for all CPMs while taking a fraction of the time and expertise. The original PROBAST can only reclassify some low ROB CPMs according to the short form to high ROB, not the other way around. In this particular sample, only a single CPM was so reclassified from low to high ROB.

Most CPMs in our study were rated high ROB, in line with previous systematic reviews using PROBAST.^16-20^ This is inherent to the structure of PROBAST: one incorrectly performed item in one domain determines an overall judgment of high ROB. While the high percentage of models with high ROB might be interpreted as a reflection of the low overall quality of the literature, it might also reflect limitations of the tool. We found substantial variation in the number of items violated by any individual CPMs, which suggests variation in methodological rigor among high ROB CPMs. Moreover, the discriminatory performance within the high ROB group varied widely (IQR of dAUC ranging from −33% to +2.6%). While the purpose of this study was to validate the ROB assessment by PROBAST, future work might explore whether it is useful to identify an “intermediate” ROB category, which might provide a more graded, less stringent assessment.

Our study has several limitations. Poor model performance at external validation can be due to the relatedness of the settings where the CPM is developed versus validated, changes in case-mix, and methodological problems (i.e., statistical overfitting).^21, 22^ Our analyses were focused on ROB caused by methodological issues, while relatedness of the validation cohort and case-mix differences will also affect the observed change in discrimination.^23, 24^ Relatedness rubrics were used to adjust for differences in relatedness of the validation cohort. The effect of case mix differences is potentially measurable through a model based concordance (c) statistic,^25, 26^ but this requires primary patient-level data to compute. Furthermore, discrimination is not the only, or even necessarily the most important metric by which to evaluate model performance. The net benefit of applying a model for decision support in a new population is a function of both discrimination and calibration.^27^ Calibration might be more sensitive to bias in model development, since it depends on the consistency of both measured and unmeasured predictor effects. However, calibration metrics are incompletely and inconsistently reported; when reported, the metrics provided are usually either clinically uninformative (e.g., the Hosmer-Lemeshow test) or difficult to quantitatively analyse (e.g., graphical).^21^ We also found that relevant information on model development was often lacking, for example about the selection of predictors, or handling of missing data. This emphasizes the importance of standardizing the reporting of CPM studies by conforming to the TRIPOD guidelines.^28, 29^

Personalized, risk-based decision making has become increasingly important in medicine, leading to a substantial increase in the number of published CPMs over the past 2 decades.^2^ Many prediction models are available, while their use in clinical practice is often limited.^30-32^ Also, the quality of reporting is considered variable and insufficient.^33-37^ Therefore, reviews of CPM studies are important to help clinicians and policy makers decide which CPM should be promoted in evidence-based guidelines or implemented in practice. The PROBAST scores, either from the full instrument or from a subset of questions, appear valid to assess and compare model quality. A recent systematic review of prediction models for COVID-19 identified 145 models that were all high ROB, mostly based on a combination of poor reporting and poor methodological conduct.^38^ Adequate sample size and a rigorous methodological approach are required to develop robust CPMs that can be applied in clinical practice.

In conclusion, high ROB is pervasive and is associated with poorer model performance at validation, supporting the application of PROBAST in reviews of CPMs. A subset of questions from PROBAST may be particularly useful for high volume assessments, when classification into high and low ROB categories is the primary goal. Furthermore, the high prevalence of high ROB models emphasizes the need to improve methodological quality of prediction research.

## Supporting information

PRISMA checklist

Supplemental Material- Short Form Guidelines

## Data Availability

We used publications from the Tufts Predictive Analytics and Comparative Effectiveness (PACE) CPM Registry

http://www.pacecpmregistry.org/

## 5.0 Acknowledgements

We thank Christine M. Lundquist, Rebecca E. H. Maunder, and Jennifer S. Lutz (research assistants at the Predictive Analytics and Comparative Effectiveness (PACE) Center, Tufts Medical Center, Boston, MA) for their help with the data extraction and assessment of the short form.

