## Supplemental Material- Short Form Guidelines for "Large-scale validation of the Prediction model Risk Of Bias ASsessment Tool (PROBAST) using a short form: high risk of bias models show poorer discrimination"

### Supplementary material

**Short form guidelines**

This short form consists of 6 PROBAST questions that can be answered with “yes”, “no”, or “no information”. Each incorrectly performed item (ie, answered with “no”) counts as 1 point, allowing a total score of 0 to 6 points. Models are categorized based on the total score, with a score of 1 or more indicating high ROB. Models with 0 points are rated low ROB or, when at least 1 item was answered with “no information”, unclear ROB.

The effect of using univariable selection and not correcting for optimism will be negligible when the effective sample size is very large. Therefore, the points for these items will be dropped when the EPV is very large (definition: EPV candidate predictors ≥25, or, when the number of candidate predictors is missing, EPV final model ≥50).

Important changes or additions to the definitions in the original PROBAST tool are marked red.

1) **Outcome assessment** – combination of PROBAST questions 3.1, 3.2, and 3.4

*Was the outcome determined appropriately in a similar way for all patients, using a standard measure or definition?*

a. **Yes:**
   1. A method of outcome determination has been used which is considered optimal or acceptable by guidelines or previous publications on the topic.

Note: This is about level of measurement error within the method of determining outcome, not about whether the definition of the outcome method is appropriate.

2. *And*, the method of outcome determination is objective *or* a standard outcome definition is used *or* pre-specified categories are used to group outcomes.
3. *And*, outcomes were defined and determined in a similar way for all participants.
b. **No:**
   1. A clearly suboptimal method has been used that causes unacceptable error in determining outcome status in participants or if a measure that involves subjective judgment or special training was used by inexperienced persons.
   2. *Or*, the outcome definition was not standard and not pre-specified.
   3. *Or*, outcomes were clearly defined and determined in a different way for some participants.

Notes & examples:

Mortality is an example of a very objective measure and will often be rated as “yes”, when determined in a similar way for all patients.

Using the modified Rankin Scale (a score ranging from 0 to 6) is considered to be an accepted outcome measure for stroke, but can still be rated as “no” when assessed by inexperienced trainees.

2) **Events per variable (EPV)** – PROBAST question 4.1

*Were there a reasonable number of participants with the outcome?*

a. **Yes:** the ratio of the number of participants with the outcome relative to the number of candidate predictors is at least 20, or relative to the number of final covariates at least 40 (EPV candidates ≥20 or EPV final model ≥40).
b. **No:** the ratio of the number of participants with the outcome relative to the number of candidate predictors is less than 10, or relative to the number of final covariates less than 20 (EPV candidates <10 or EPV final model <20).
c. **No information:**
   1. There is no information on the number of (candidate) predictor parameters or number of participants with the outcome, such that the EPV cannot be calculated.
   2. *Or*, the ratio of the number of participants with the outcome relative to the number of candidate predictors is between 10 and 20, or relative to the number of final covariates between 20 and 40 (EPV candidates 10-20 or EPV final model 20-40).

Notes & examples:

Only when the number of candidate predictors is unclear, the number of items reported in the univariable analysis should be used. If that is not reported, than the number of parameters in the final model.

Candidate variables = all variables that were used in the univariable or multivariable selection process. To calculate the EPV, use the number of candidate predictor parameters: continuous variable = 1 parameter, non-linear variable (eg, exponential, fractional polynomial or spline) = 2 parameters, categorical variable = number of categories minus 1.

Examples of the number of candidate predictors:

1. Age (continuous) = 1 parameter
2. Gender (male/female) = 1 parameter
3. Blood pressure (odds ratio for every mmHg <120 & odds ratio for every mmHg >120) = non-linear = 2 parameters
4. Location of occlusion (M1/M2/ICA/other) = 4 categories = 3 parameters
3) **Continuous predictors** – PROBAST question 4.2

*Were continuous and categorical predictors handled appropriately?*

a. **Yes:**
   1. Continuous predictors are not converted into ≥2 categories (ie, dichotomized or categorized) when included in the model.
   2. *Or,* continuous predictors are converted into ≥2 categories based on pre-specified or widely accepted cut points (this should be explicitly mentioned in the paper or statistical protocol).
   3. *Or,* continuous predictors are examined for non-linearity (eg, with exponential, fractional polynomial or spline function).
   4. *Or,* categorical predictor groups are defined using a pre-specified or widely accepted method (ie, not based on the own data).
b. **No:** continuous predictors are converted into ≥2 categories *or* categorical predictor groups definitions are based on the “optimal” cut point or threshold in the current dataset.
c. **No information:**
   1. Continuous predictors are converted into ≥2 categories but it is not mentioned how these cut points were chosen.
   2. *Or*, it is unclear how categorical predictor groups were defined.

Notes & examples:

Dichotomizing means that a variable with continuous values is split in 2 or more groups (eg, age <65 and age ≥65), which causes loss of efficiency. Is can also cause risk of bias when the cut points are based on the current dataset.

Pre-specified = cut points or categories that were determined in the papers methods section or in a previously published paper.

If the cut points themselves are not specified but it is specified how to get the cut point (eg, use the median), it can be rated “yes” (it may not be good modeling practice, but it should not lead to high risk of bias).

4) **Missing data** – PROBAST question 4.4

*Were participants with missing data handled appropriately?*

a. **Yes:**
   1. The study explicitly reports that there are no missing values of predictors or outcomes.
   2. *Or*, missing values are handled using multiple imputation.
   3. *Or,* <5% of all patients were excluded based on missing values.
   4. *Or,* a clear justification was declared for the method of missing data handling.
b. **No:**
   1. >5% or an unknown percentage of participants with missing data are omitted from the analysis.
   2. *Or*, the method of handling missing data is clearly flawed (eg, single imputation, missing indicator method or inappropriate use of last value carried forward).
   3. *Or,* the study had no explicit mention of methods to handle missing data.

Notes & examples:

If there is no explicit mention missing data or the methods to handle missing data, this question should be answered as “no” and not as “no information”.

Adding missing values to the reference class of a categorical variable can only be rated appropriate (ie, “yes”) when at least 80% of the patients are in the reference class and <10% of the values are missing.

Using a separate category for missing values of a categorical variable is only appropriate when the model is applied to a similar setting (ie, when the pattern of missingness and the cause of missing values in the validation cohort will be similar as in the derivation cohort), but in general it should be rated as “no”.

5) **Univariable analysis** – PROBAST question 4.5

*Was selection of predictor based on univariable analysis avoided?*

a. **Yes:** the predictors are *not* selected based on univariable analysis prior to multivariable modeling (ie, predictors in the multivariable analysis were pre-specified or chosen based on expert opinion).
b. **No:** the predictors are selected based on univariable analysis prior to multivariable modelling (ie, predictors were first analyzed one at a time and only included in the multivariable model if this analysis was significant).
c. **Unclear:** there is insufficient information to indicate that univariable selection is avoided.

Notes & examples:

If there is no evidence of univariable selection, the question should be answered “yes” (ie, the authors don’t have to mention specifically that they didn’t use it).

If univariable analysis was performed, but the authors explicitly mention that predictors were included in the multivariable analysis regardless of these results, it should be rated “yes”.

If univariable analysis was performed, but it is not clear whether this affected predictor selection for the multivariable model, it should be rated “no information”.

Exception: if it is explicitly mentioned that the internal validation process included all variable selection procedures, this question can be answered with “yes”.

6) **Correction for overfitting/optimism** – PROBAST question 4.8

*Was model overfitting and optimism in model performance accounted for?*

a. **Yes:** internal validation techniques, such as bootstrapping and cross-validation, have been used to account for any optimism in model fitting, *and* subsequent adjustment of the prediction model performance measures have been applied (ie, “internally validated AUC”).
b. **No:**
   1. No internal validation has been performed.
   2. *Or*, internal validation consisted only of a single random split-sample of participant data.
   3. *Or,* the bootstrapping or cross-validation did not include all model development procedures including any variable selection.

Notes & examples

Internal validation and adjustment of model performance measures can be indicated in the text with for example: “bootstrapping”, “optimism correction”, “lasso”, “penalized regression”, or “shrinkage”. A random split sample is not considered a valid internal validation technique.

If there is nothing reported about internal validation or correction for overfitting/optimism, we assume that it has not been performed (so this item should be rated “no”).

External validation is not integrated in the PROBAST short form. Only internal validation is used to correct the apparent performance measure in the development cohort (ie, derivation AUC) for optimism, and can thereby reduce bias of the reported AUC.
