## Supplemental Table 1 for "Large-scale validation of the Prediction model Risk Of Bias ASsessment Tool (PROBAST) using a short form: high risk of bias models show poorer discrimination"

### Supplementary material

**Table S1.** Interrater agreement in the assessment of PROBAST items before the final consensus meeting.

|  | **Percentage agreement** | **Kappa** |
| --- | --- | --- |
| **1. Participants** | | |
| 1.1 Were appropriate data sources used, e.g., cohort, RCT, or nested case–control study data? | 93% | 0.34 |
| 1.2 Were all inclusions and exclusions of participants appropriate? | 76% | 0.45 |
| **2. Predictors** | | |
| 2.1 Were predictors defined and assessed in a similar way for all participants? | 91% | 0.28 |
| 2.2 Were predictor assessments made without knowledge of outcome data? | 85% | 0.67 |
| 2.3 Are all predictors available at the time the model is intended to be used? | 91% | 0.50 |
| **3. Outcome** | | |
| 3.1 Was the outcome determined appropriately? | 94% | 0.42 |
| 3.2 Was a prespecified or standard outcome definition used? | 90% | 0.05 |
| 3.3 Were predictors excluded from the outcome definition? | 97% | 0 |
| 3.4 Was the outcome defined and determined in a similar way for all participants? | 80% | 0.12 |
| 3.5 Was the outcome determined without knowledge of predictor information? | 74% | 0.57 |
| 3.6 Was the time interval between predictor assessment and outcome determination appropriate? | 88% | -0.05 |
| **4. Analysis** | | |
| 4.1 Were there a reasonable number of participants with the outcome? | 92% | 0.88 |
| 4.2 Were continuous and categorical predictors handled appropriately? | 75% | 0.52 |
| 4.3 Were all enrolled participants included in the analysis? | 75% | 0.46 |
| 4.4 Were participants with missing data handled appropriately? | 82% | 0.68 |
| 4.5 Was selection of predictors based on univariable analysis avoided? | 92% | 0.83 |
| 4.6 Were complexities in the data (e.g., censoring, competing risks, sampling of control participants) accounted for appropriately? | 49% | 0.19 |
| 4.7 Were relevant model performance measures evaluated  appropriately? | 84% | 0.84 |
| 4.8 Were model overfitting and optimism in model performance accounted for? | 79% | 0.55 |
| 4.9 Do predictors and their assigned weights in the final model correspond to the results from the reported multivariable analysis? | 82% | 0.74 |
| **Total ROB** | **90%** | **0.33** |
