## Supplemental Table 2 for "Large-scale validation of the Prediction model Risk Of Bias ASsessment Tool (PROBAST) using a short form: high risk of bias models show poorer discrimination"

### Supplementary material

**Table S2.** Example of derivation-validation pairs with a 17% difference in delta AUC.

| **Derivation AUC** | **Short form classification** | **Delta AUC** | **Validation AUC** | **Absolute difference in validation AUC** |
| --- | --- | --- | --- | --- |
| 0.90 | High ROB | -10% | 0.86 | 0.07 |
|  | Low ROB | +7% | 0.93 |  |
| 0.90 | High ROB | -25% | 0.80 | 0.07 |
|  | Low ROB | -8% | 0.87 |  |
| 0.80 | High ROB | -10% | 0.77 | 0.05 |
|  | Low ROB | +7% | 0.82 |  |
| 0.80 | High ROB | -25% | 0.73 | 0.05 |
|  | Low ROB | -8% | 0.78 |  |
| 0.70 | High ROB | -10% | 0.68 | 0.03 |
|  | Low ROB | +7% | 0.71 |  |
| 0.70 | High ROB | -25% | 0.65 | 0.03 |
|  | Low ROB | -8% | 0.68 |  |
| 0.60 | High ROB | -10% | 0.59 | 0.02 |
|  | Low ROB | +7% | 0.61 |  |
| 0.60 | High ROB | -25% | 0.57 | 0.02 |
|  | Low ROB | -8% | 0.59 |  |
